# Clinical and economic inpatient burden of respiratory syncytial virus (RSV) infections in children < 2 years of age in Germany, 2014-2019: a retrospective health claims analysis

**DOI:** 10.1101/2024.02.12.24302675

**Authors:** Caroline Lade, Lea Bayer, Bennet Huebbe, Jennifer Riedel, Sima Melnik, Gordon Brestrich, Christof von Eiff, Tobias Tenenbaum

## Abstract

**Background:** Respiratory syntactical virus (RSV) is a common cause for severe lower respiratory tract infections (LRTI) in children <2 years of age in Germany – though little is known about the clinical and economic burden of RSV in children with and without risk factors per month of life.

**Methods:** In a retrospective health claims analysis, we identified RSV inpatient cases between 2014 and 2019. We assessed incidence rates, mortality rate, health resource utilization, associated direct costs per case and excess costs for 30, 90 and 365 days after hospital admission matched to a control group. The outcomes are reported separately for the first and second year of life (i.e., for infants and toddlers) and were stratified by month of life, preterm and risk status (i.e., presence of underlying disease: chronic respiratory or cardiac disease, immunosuppression, neurological diseases, diabetes, conditions originating in the perinatal period).

**Results:** RSV-attributable hospital incidence rate was higher in infants (30.25/1,000) than toddlers (14.52/1,000), highest in the first three months of life (44.21/1,000), in infants born preterm (64.76/1,000) or with any underlying disease (54.85/1,000). Mortality rate was also higher for infants (0.08/1,000) than toddlers (0.04/1,000). Mean 30-day excess costs ranged from 2,953 € for infants born full-term at no risk, hospitalized for 5 days, to 6,694 € for infants born extremely premature, hospitalized for 7 days.

**Conclusion:** In Germany, the clinical and economic burden of RSV is substantial, especially in the most vulnerable population, that is, very young infants, those born premature and/or those with an underlying disease.

## Introduction

Respiratory syncytial virus (RSV) is a common cause of acute lower respiratory tract infections (LRTI) at all ages worldwide. RSV infections are most severe, often leading to hospitalizations, in children under the age of 5 [1]. In children, RSV manifests with symptoms ranging from rhinitis, bronchiolitis to pneumonia [2]. By the age of two years, most children have had an RSV infection at least once [3]. Severe cases occur especially in high-risk groups which include children born prematurely and children with underlying diseases like chronic respiratory or cardiac disease, immunosuppression, neurological diseases, diabetes, or conditions originating in the perinatal period [4]. Morbidity and mortality due to RSV are higher in children at risk than no risk [5]. However, little is known about either clinical or economic burden of RSV in the most vulnerable children < 2 years of age in Germany.

The global burden of RSV was estimated in 2019 at 33 million episodes a year among children under five years of age or younger, with 3.6 million hospitalizations, 26,300 RSV in-hospital deaths, and 101,400 RSV-attributable overall deaths [6]. The majority of cases were accounted for by children under one year of age. For industrialized countries, hospital incidence rates were reported for RSV-associated acute LRTI at 38.5/1,000 in children under one year of age and 24.3/1,000 in children younger than 5 years of age [6]. Hospital incidence rates are highest within the first month of age, decreasing steadily month by month thereafter [7, 8].

In Germany, the clinical burden of RSV in children < 2 years of age has only very recently been described [9–11] but data on the burden of RSV in more vulnerable children are still missing, that is for very young infants, children born prematurely or with an underlying disease. Hospital incidence rates of 14.9-28.6 per 1,000 infants (i.e., first year of life) were reported between 2019-2022 [11]. For toddlers (i.e., second year of life), the hospital incidence rate ranged between 2.5-5.6 per 1,000 persons. Thereby, most RSV hospital cases were identified in infants under six months of life based on medical records of children under five years of age between 2015 and 2018 [10] – but no incidence rates were reported. Despite high awareness and good testing practice, incidence rates in children may be underestimated [12]. Likely large proportions of cases of respiratory diseases that are not readily identified as RSV may be documented as RSV-attributable unspecified diagnoses of, for instance, unspecified bronchiolitis, in young children under the age of 2 years [13], contribute to an underreporting of RSV. Such underestimation in combination with an ever-increasing RSV hospital incidence rate that almost doubled since 2010 [9] highlights the importance to better understand the clinical burden of RSV in more detail, especially in the most vulnerable population of children in Germany.

In addition to the clinical burden, RSV infections also pose a substantial economic burden to the healthcare system. A recent review summarizing data from Europe, North America, and Australia between 1987 and 2017 reported average costs of 3,452 € per inpatient episode among children under five years of age [14]. Specifically for Germany, costs of 3,001 € - 3,961 € were reported for RSV cases in children ≤2 years of age between 2019-2022 [11]. For children under one year of age, costs of 3,329 € were reported between 2015-2018 [9]. Costs increased to 7,825 € - 11,045 € for severe cases that required ICU admission [11]. Since previous reports of costs in 2002, costs have increased by approximately 15% [15]. With the highest clinical burden in infants with risk factors, more detailed data are needed on healthcare resource utilization (HRU) and associated costs of RSV in the most vulnerable population to better understand the economic burden in Germany.

To fill in these gaps in the clinical and economic burden of RSV in children in Germany, we conducted retrospective analyses using health claims data to assess the incidence rate, mortality, HRU, and costs associated with RSV in hospitalized children <2 years of age in Germany from 2014 to 2019, stratified by month of life and the presence of risk factors (prematurity and/or underlying disease).

## Methods

### Study design and data source

In this study, we conducted an observational, retrospective database analysis. Anonymized health claims data were obtained from the German Analysis Database for Evaluation and Health Services Research [*Deutsche Analysedatenbank für Evaluation und Versorgungsforschung*] (DADB) administered by Gesundheitsforen Leipzig GmbH [16] containing Statutory Health Insurance (SHI) data on approximately four million insured persons (∼5 % of the SHI population), representative of the German population in terms of age, sex and morbidity.

### Study population and case identification

Health claims data were selected from insured children ≤ 24 months of life, born between January 1^st^, 2013, and December 31^st^, 2019, who were fully observable for either their first or second year of life or until death. Cases were identified within an observation period between January 1^st^, 2014 and December 31^st^, 2019, to avoid confounds due to irregular RSV seasons during and after the COVID-19 pandemic [17]. Case identification was based on either a narrow definition including RSV-specific ICD-10-GM codes, J12.1 (acute pneumonia due to RSV), J20.5 (acute bronchitis due to RSV), J21.0 (acute bronchiolitis due to RSV), and B97.4 (RSV classified elsewhere) or a wide definition including both RSV-specific diagnosis codes and RSV-attributable ICD-10-GM codes of unspecified bronchiolitis (J21.8, J21.9), bronchitis (J20.8, J20.9), pneumonia (J12.8, J12.9, J18.8, J18.9) or acute lower respiratory tract infections (J22, UBP). In this wide definition, UBP cases were included if the diagnosis occurred during the RSV season as defined by RKI for each year, typically between December and February according to Cai et al. [18] (see Supplemental Table 1). Previous studies have repeatedly employed such wide case definition [e.g., 19, 20] to increase the sensitivity of capturing RSV cases without a substantial loss of specificity [21]. For case identification, main and secondary diagnoses were taken into account. All analyses were carried out separately for incident infants (first year of life) and toddlers (second year of life; for case characteristics, see Table 1). The age of patients in months of life was estimated at the time of the index diagnosis. For the analysis of costs, first, patients with multiple cases (n = 824 cases, 10.7 %), second, patients who were not fully observable for the maximum follow-up period of one year after the index date (n = 208 cases, 3% ), third, patients for who there was no exact match amongst control patients (n = 25, 0.38 %) were excluded in order to reliably and consistently assess long-term costs (Figure S1). Patients were further characterized by preterm status (full-term, extreme preterm and early/late preterm) and risk status (no risk, any risk, Table 1). Preterm status was defined by the presence of the ICD-10 codes P07.2, which characterizes newborns with extreme prematurity, born before the 29^th^ gestational week (henceforth extreme preterm), and P07.3 which characterizes other preterm infants, infants born between the 29^th^ and the 37^th^ gestational week (henceforth early/late preterm). Risk status was defined by the presence of at least one risk factor based on ICD-10 codes for conditions originating in the perinatal period, chronic or congenital cardiac or respiratory disease, immunosuppression, neurological disease, diabetes mellitus or a patient history of prophylactic immunotherapy (Table S2). As conditions originating in the perinatal period also include prematurity, the clinical burden of RSV was additionally described for children at risk excluding prematurity, that is for children with underlying disease born full-term.

**Table 1.** Number (n) and characteristics of inpatient cases between 2014-2019 in infants and between 2015-2019 in toddlers.

| Characteristic | Infants |  | Toddlers |  |
| --- | --- | --- | --- | --- |
|  | 1-12 months |  | 13-24 months |  |
|  | Incidence | HRU/costs | Incidence | HRU/costs |
|  | cohort,<br>n = 5,565 | cohort,<br>n = 4,909 | cohort,<br>n = 2,131 | cohort,<br>n = 1,730 |
|  | n (%) | n (%) | n (%) | n (%) |
| **Age in months** (Mean, ±SD) | 5.4 (±3.2) | 5.2 (±3.2) | 17.9 (±3.3) | 17.8 (±3.3) |
| **Sex (female)** | 2,265 (40.7) | 2,033 (41.4) | 888 (41.7) | 723 (41.8) |
| **Episode type** |  |  |  |  |
| RSV | 3,522 (63.3) | 3,223 (65.7) | 679 (31.9) | 573 (33.1) |
| UBP | 1,859 (33.4) | 1,538 (31.3) | 1,390 (65.2) | 1,112 (64.3) |
| RSV+UBP | 184 (3.3) | 148 (3.0) | 62 (2.9) | 45 (2.6) |
| **Full term status** |  |  |  |  |
| full term | 4,825 (86.7) | 4,313 (87.9) | 1,815 (85.2) | 1,505 (87.0) |
| preterm | 670 (12.0) | 559 (11.4) | 272 (12.8) | 208 (12.0) |
| extreme preterm | 70 (1.3) | 37 (0.8) | 44 (2.1) | 17 (1.0) |
| **Risk factors** |  |  |  |  |
| any risk factor | 1,714 (30.8) | 1,430 (29.1) | 748 (35.1) | 547 (31.6) |
| no risk factor | 3,851 (69.2) | 3,479 (70.9) | 1,383 (64.9) | 1,183 (68.4) |
| **ICU stay** |  |  |  |  |
| ICU stay | N/A | 43 (0.9) | N/A | 12 (0.7) |
| no ICU stay | N/A | 4,866 (99.1) | N/A | 1,718 (99.3) |
| **Ventilatory support** |  |  |  |  |
| ventilatory support | N/A | 288 (5.9) | N/A | 18 (1.0) |
| no ventilatory support | N/A | 4,621 (94.1) | N/A | 1,712 (99.0) |

#### Definition of episodes

Hospital episodes were defined as overnight stays. The start of an episode was defined by the admission date of either main or secondary RSV or UBP diagnosis (i.e., index date). An episode lasted until the discharge date or until no further RSV or UBP diagnosis occurred within 30 days of the previous diagnosis to capture possible readmissions [10] and to avoid an overestimation of case counts. Two types of episodes were classified: first, RSV episodes which were based on either RSV-specific diagnosis codes only or RSV-specific diagnosis codes plus an additional UBP diagnosis code during the duration of the episode; second, UBP episodes that are based on UBP diagnosis codes only. For the wide case definition, both RSV and UBP episodes were considered for the assessment of outcomes measures; for the narrow definition only RSV episodes were considered.

#### Assessment

Study outcome measures included incidence rates, 90-day all-cause mortality rate, all-cause healthcare costs and HRU. Incidence rates were calculated as number of cases divided by the annual number of newborns in the DADB (Table S6) and are expressed per 1,000 persons per year. For the estimation of incidence rates per month of life, the annual base population was divided by 12 months. For age group stratifications, the annual base population was divided by 12 and then multiplied by the number of months the respective age group encompasses, assuming approximately equal numbers of children per month of life.

The 90-day all-cause mortality rate was calculated as the number of deceased patients within the episode plus 90 days thereafter divided by the total population size across the observation period. Due to relatively low numbers of fatal cases, mortality rates could only be computed across the entire study period from 2014-2019. The mortality rate was stratified by preterm and risk status.

HRU assessment included hospital length of stay (LOS), ICU admission status and ventilation use. LOS was assessed per episode and stratified by <5, 5-8 and >8 days. In addition, LOS was assessed for all-cause hospitalizations during 30, 90 and 365 days after the index date. ICU stay was assessed per episode based on admission to or discharge from ICU. Ventilation use was assessed per episode based on OPS codes for oxygen therapy (OPS 8-720), non-invasive ventilatory support (High Flow Nasal Cannula, HFNC: OPS 8-711.4, OPS 8-712.1; Continuous Positive Airway Pressure, CPAP: OPS 8-711.0, OPS 8-712.0), invasive mechanical ventilation (OPS 8-711.1, OPS 8-711.2).

Mean direct healthcare costs were calculated per episode, mean excess costs were estimated for short- and longer-term follow-up time windows (30, 90, 365 days after hospital admission). Excess costs were compared 1:1 to matched control patients across healthcare sectors based on month of life, sex, preterm status using Mann-Whitney-Wilcoxon test for continuous variables. P-values were adjusted for multiple comparisons, if necessary, using the Benjamini-Hochberg procedure to limit the probability of false detections. The control cohort was also observable for at least one year after the index date of the matched incident patient and did not have an RSV diagnosis within that time or a UBP diagnosis during the RSV season. 25 cases (0.4%) for which there was no exact match were excluded from all cost analyses. All cost values were adjusted to 2019 per consumer price index [22] per case per time window before being averaged for each time window across the study period 2014-2019.

All outcomes were stratified, whenever case numbers allowed, by episode type (RSV-UBP, RSV, UBP), month of life (1-12 months and 13-24 months), calendar month (January to December), preterm (full-term, extreme preterm and early/late preterm) and risk status (no risk, any risk). All analyses were descriptive and exploratory. As there were no a priori hypotheses, no planned comparisons were performed.

Finally, we extrapolated mean costs to the population-level in order to estimate the average annual economic burden for Germany. To this end, we first determined an extrapolation factor per calendar year based on the number of live births per year (mean across 2014 - 2019 = 765,680 live births; [23]) and the number of newborns in the DADB (both in Table S6) and then multiplied that with the annual case numbers. The extrapolated mean annual number of hospitalizations were multiplied with the mean excess costs for 30 days after hospital admission.

*Ethical considerations.* This study used aggregated and anonymized health claims data, therefore not requiring approval from Institutional Review Boards or Ethical Committees or informed consents from patients. The study was conducted in accordance with legal and regulatory requirements and research practices described in the “Good Epidemiological Practice guidelines issued by the International Epidemiological Association” [24].

## Results

Between 2014-2019, there were 183,952 infants within their first year of life and 146,752 toddlers in their second year of life in the database (for more information on the annual base population, see Table S6).

### Clinical burden – incidence and mortality

We identified 5,565 cases in infants (based on 5,248 main diagnoses, 317 secondary diagnoses) and 2,131 cases in toddlers (based on 1,927 main diagnoses and 204 secondary diagnoses) across the study period (for further characteristics see Table 1; also see patient flow in Figure S1). Most cases were identified within the RSV season between December and March (88% of the cases in infants, 93% in toddlers). Among infants, 66.6% of cases were identified based on an RSV diagnosis code; in toddlers it was 34.8%. In infants, 45.65% cases were diagnosed with acute bronchiolitis due to RSV, 29.86% with acute bronchitis due to RSV, 21.70% with acute pneumonia due to RSV and 2.79% with RSV classified elsewhere. In contrast, in toddlers, 43.10 % cases were diagnosed with acute pneumonia due to RSV, 32.90% with acute bronchitis due to RSV, 18.97% with acute bronchiolitis due to RSV and 5.03% with RSV classified elsewhere. Moreover, 86.70% of infants were born full-term, 13.30% preterm, similar in toddlers. 30.8% of infants and 35.1% of toddlers were considered at risk (see Table 1 for more specific case characteristics).

The hospitalization incidence rate in infants was estimated at 28.95-33.07 per 1000 between 2014 and 2019, with a mean of 30.25 per 1,000 (Table 2). In toddlers, the incidence rate was estimated at 10.95-18.13 per 1,000 between 2015 and 2019, with a mean of 14.52 per 1,000 (for toddlers we only included data from 2015 onwards to fully observe all children in the database within their second year of life who were born between 2013-2019. More specifically, for infant cases based on an RSV-specific diagnosis, the annual hospitalization incidence rate was 17.25-22.89 per 1,000 between 2014-2019 with a mean of 20.15 per 1,000; for toddlers, it was 3.70-6.58 per 1,000 with a mean of 5.05 per 1,000. Extrapolated to the population-level, that is 23,152 cases of infants (15,469 RSV-specific cases) and 11,228 cases of toddlers (3,907 RSV-specific cases) per year.

**Table 2.** Case Numbers (N) and Hospital Incidence Rates (IR) per 1,000 Person-Years per Calendar Year in Children ≤ 2 years of age between 2014-2019.

| Month<br>of life | 2014 |  | 2015 |  | 2016 |  | 2017 |  | 2018 |  | 2019 |  | 2014<br>–<br>2019 |  |
| --- | --- | --- | --- | --- | --- | --- | --- | --- | --- | --- | --- | --- | --- | --- |
|  | N | IR | N | IR | N | IR | N | IR | N | IR | N | IR | N | IR |
| *RSV-UBP* |  |  |  |  |  |  |  |  |  |  |  |  |  |  |
| 1-3 | 319 | 41.69 | 315 | 41.71 | 334 | 43.30 | 375 | 47.45 | 324 | 41.55 | 366 | 49.66 | 2,033 | 44.21 |
| 4-6 | 290 | 37.90 | 262 | 34.69 | 288 | 37.34 | 275 | 34.80 | 240 | 30.78 | 271 | 36.77 | 1,626 | 35.36 |
| 7-12 | 403 | 26.34 | 299 | 19.79 | 330 | 21.39 | 273 | 17.27 | 339 | 21.74 | 262 | 17.77 | 1,906 | 20.72 |
| 1-6 | 609 | 39.80 | 577 | 38.20 | 622 | 40.32 | 650 | 41.13 | 564 | 36.16 | 637 | 43.21 | 3,659 | 39.78 |
| 1-12 | 1,012 | 33.07 | 876 | 29.00 | 952 | 30.86 | 923 | 29.20 | 903 | 28.95 | 899 | 30.49 | 5,565 | 30.25 |
| 13-24 | NA | NA | 399 | 13.80 | 521 | 18.13 | 323 | 10.95 | 463 | 15.29 | 425 | 14.49 | 2,131 | 14.52 |
| 1-24 | NA | NA | 1,275 | 21.57 | 1,473 | 24.72 | 1,246 | 20.39 | 1,366 | 22.22 | 1,324 | 22.51 | 7,696 | 23.27 |
| *RSV-specific* |  |  |  |  |  |  |  |  |  |  |  |  |  |  |
| 1-3 | 237 | 30.98 | 243 | 32.17 | 260 | 33.71 | 313 | 39.61 | 264 | 33.86 | 318 | 43.14 | 1,635 | 35.55 |
| 4-6 | 174 | 22.74 | 169 | 22.38 | 204 | 26.45 | 214 | 27.08 | 158 | 20.26 | 213 | 28.90 | 1,132 | 24.62 |
| 7-12 | 156 | 10.19 | 147 | 9.73 | 146 | 9.46 | 147 | 9.30 | 174 | 11.16 | 169 | 11.46 | 939 | 10.21 |
| 1-6 | 411 | 26.86 | 412 | 27.27 | 464 | 30.08 | 527 | 33.34 | 422 | 27.06 | 531 | 36.02 | 2,767 | 30.08 |
| 1-12 | 567 | 18.53 | 559 | 18.50 | 610 | 19.77 | 674 | 21.32 | 596 | 19.11 | 700 | 23.74 | 3,706 | 20.15 |
| 13-24 | NA | NA | 115 | 3.98 | 147 | 5.12 | 133 | 4.51 | 137 | 4.53 | 204 | 6.95 | 741 | 5.05 |
| 1-24 | NA | NA | 674 | 11.40 | 757 | 12.70 | 807 | 13.20 | 733 | 11.93 | 904 | 15.37 | 4,447 | 13.45 |

Across the first two years of life, 50% of cases occur already within the first 6 months of life, with specifically high incidence rates in the second and third month of life (57.28 and 44.88 per 1000 person-years, respectively), continuously decreasing thereafter, across the second year of life (Figure 1, Table S4). This is driven by infants with an RSV-specific diagnosis for which the incidence rate in the second and third month was at 45.21 and 34.84, respectively. For infants with an RSV-specific diagnosis, the incidence rate is 2.4 times higher in the first three months than for the remaining first year of life, and 2.9 times higher in the first 6 months than in the second half a year of life.

**Figure 1.**
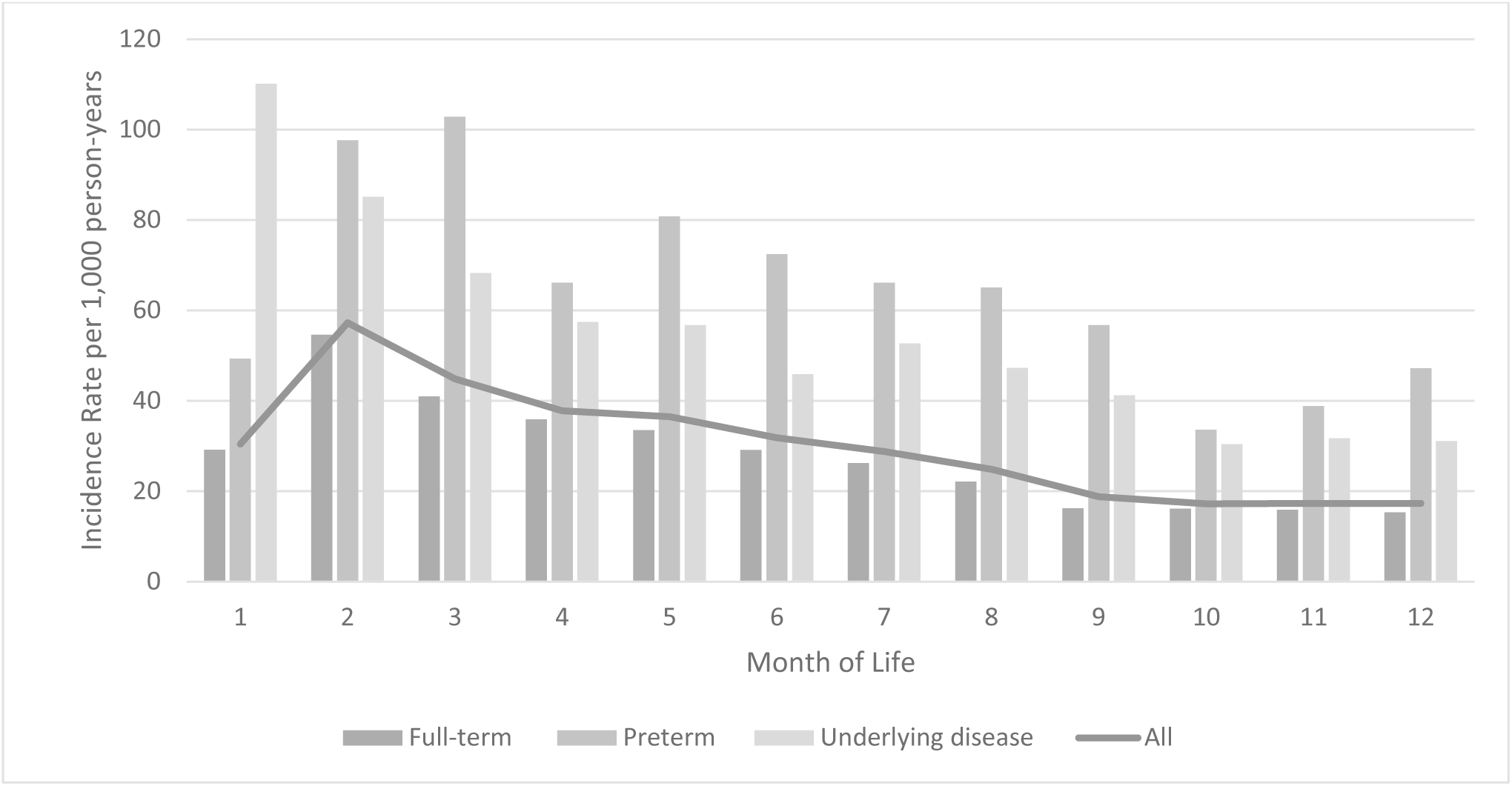
Hospital Incidence Rate per cross stratified by months of life and preterm status, risk status 2014-2019

Incidence rates were highest for infants born prematurely (preterm: 64.76, full-term: 27.97) and for infants at risk including prematurity (any risk: 58.73, no risk: 28.92; for more details see Table 3), especially shortly after birth (Figure 1, Table S4). Compared to infants at no risk, incidence rates were 2.2 times higher in infants with any underlying disease, 2.5 times higher in early/late preterms, and 3.3 times higher in infants born extremely premature. For toddlers, prematurity and underlying risk factors remain to play a great role with highest incidence rates in toddlers that were born prematurely (preterm: 34.49, full-term: 13.19) and those at risk (at risk: 28.92, no risk: 11.44). Compared to toddlers at no risk, incidence rates were 2.3 times higher in toddlers with any underlying disease, 2.8 times higher in toddlers with early/late prematurity, and 6.1 times higher in toddlers with extreme prematurity.

**Table 3.** Case Numbers (N) and Hospital Incidence Rates (IR) per 1000 per person-years between 2014-2019 stratified by Preterm and Risk Status.

|  | Infants |  |  | Toddlers |  |  |
| --- | --- | --- | --- | --- | --- | --- |
|  | DADB* | N | IR | DADB* | N | IR |
| *RSV-UBP* |  |  |  |  |  |  |
| No risk | 154,769 | 3,851 | 24.88 | 120,892 | 1,383 | 11.44 |
| Any risk** | 29,183 | 1,714 | 58.73 | 25,860 | 748 | 28.92 |
| Underlying disease*** | 17,756 | 974 | 54.85 | 16,698 | 432 | 25.87 |
| Full-term | 172,525 | 4,825 | 27.97 | 137,590 | 1,815 | 13.19 |
| Any preterm | 11,427 | 740 | 64.76 | 9,162 | 316 | 34.49 |
| Early/late preterm | 10,583 | 670 | 63.31 | 8,530 | 272 | 31.89 |
| Extreme preterm | 844 | 70 | 82.94 | 632 | 44 | 69.62 |
| *RSV-specific* |  |  |  |  |  |  |
| No risk | 154,769 | 2,577 | 16.65 | 120,892 | 459 | 3.80 |
| Any risk** | 29,183 | 1,129 | 38.69 | 25,860 | 282 | 10.90 |
| Underlying disease | 17,756 | 646 | 36.38 | 16,698 | 166 | 9.94 |
| Full-term | 172,525 | 3,223 | 18.68 | 137,590 | 625 | 4.54 |
| Any preterm | 11,427 | 483 | 42.27 | 9,162 | 116 | 12.66 |
| Early/late preterm | 10,583 | 451 | 42.62 | 8,530 | 96 | 11.25 |
| Extreme preterm | 844 | 32 | 37.91 | 632 | 20 | 31.65 |

*Note: \* DADB base population, \*\* presence of any risk factor including any underlying disease and prematurity; \*\*\* excluding premature cases*

Between 2014-2019, 20 children died within the episode or 90 days thereafter – 14 infants and 6 toddlers. 9 of the fatal cases were born prematurely, 11 full-term. All 20 fatal cases had at least one risk factor (including prematurity). 11 children died after an RSV-specific diagnosis, 9 after a UBP diagnosis (that occurred within the RSV season). Extrapolated to the population-level, there were a total of 538 fatal cases per year, 350 infants and 188 toddlers. The annual all-cause mortality rate after an RSV infection in children under two years is estimated at 0.06 per 1,000 children, 0.08 per 1,000 infants and 0.04 per 1,000 toddlers.

### Economic burden and healthcare resource utilization related to RSV infections

Mean direct and mean excess costs as well as HRU were estimated based on 4,909 cases in infants and 1,730 cases in toddlers across the study period for which there was a 1:1 exact matched control patient and which were observable for at least one year after the index date (for further characteristics see Table 1; also see patient flow in Figure S1). Among infants, 68.7% of cases were identified based on an RSV-specific diagnosis code, in toddlers it was 35.7%.

Infants were hospitalized for a median LOS of 5 days, toddlers for 4 days, within 30 days after hospital admission. The LOS for both infants and toddlers with an RSV-specific diagnosis was 5 days. Infants and toddlers with acute pneumonia due to RSV (J12.1) had the longest median LOS of 7 and 6 days, respectively. Children with acute bronchiolitis due to RSV (J21.0) had shorter median LOS of 5 days; children with acute bronchitis due to RSV (J20.5) or RSV classified elsewhere (B97.4) had the shortest median LOS of 4 days. For infants born early/late preterm, the median LOS was 6 days. For those born extreme premature, it was 7 days or even 8 days for infant cases with an RSV-specific diagnosis. In toddlers, this was 5 days for those born early/late preterm and 6 days for those with extreme prematurity.

0.9% of cases in infants were admitted to ICU, whereby 83.7% infants were born full-term and 60.5% were between the age of 1-3 months of life. For toddlers, 0.7% of cases were admitted to ICU, whereby 91.7% were born full-term and 66.7% were between 13-18 months of life. Risk factors (including prematurity) did not increase the likelihood of ICU admission (55.8% infants, 44.7% toddlers were admitted to ICU). Further, 5.9% of infants required some form of ventilation use: 1.8% oxygen therapy, 3.6% other non-invasive ventilatory support (1.9% HFNC, 1.7% CPAP) and 0.5% invasive mechanical ventilation. Only 1% of toddlers required ventilatory support (0.9% HFNC, 0.1% CPAP).

Mean direct costs attributed to those healthcare resource utilizations were higher in infants (3,775 €) than in toddlers (3,062 €, for more details per age group see Table 4), higher for those born premature or at risk compared to children at no risk (Table 5). For infants, mean costs were 4.7 times higher for cases that required ICU admission (17,273 €) than for cases which did not require ICU admission (3,655 €). For toddlers, mean costs were even 9.5 times higher for cases that required ICU admission (27,605 €) than for cases which did not require ICU admission (2,891 €). Infant cases that required invasive mechanical ventilation support were especially high, with mean costs up to 75,188 €. For both infants and toddlers, mean costs were higher for cases with an RSV episode (4,021 € and 3,641 €, respectively) than for cases with a UBP episode (3,234 € and 2,741 €, respectively). Thereby, mean costs varied between different RSV-specific diagnosis codes. That is, in infants, in the order from high to low case numbers, on average, acute bronchiolitis due to RSV (J21.0) cost 4,061€; acute bronchitis due to RSV (J20.5) cost 2,883 €; acute pneumonia due to RSV (J12.1) cost 5,773 €; cases with RSV classified elsewhere (B97.4) cost 4,996 €. In toddlers, acute pneumonia due to RSV (J12.1) cost 4,458 €; acute bronchitis due to RSV (J20.5) cost 3,118 €; acute bronchiolitis due to RSV (J21.0) cost 3,310 €; cases with RSV classified elsewhere (B97.4) cost 3,706 €.

**Table 4.** Case Numbers (N), Mean (M) and Standard Deviation (SD) of Direct Costs and Mean Excess Costs (in €) in Infants and Toddlers between 2014 – 2019.

|  | Month<br>of life | N | Direct Costs | Excess costs |  |  |
| --- | --- | --- | --- | --- | --- | --- |
|  |  |  | Episode | 30 days | 90 days | 365 days |
|  |  |  | M (SD) | M | M | M |
| RSV-UBP |  |  |  |  |  |  |
|  | 1-3 | 1,891 | 4,738 (11,276) | 4,028 | 4,612 | 5,065 |
|  | 4-6 | 1,430 | 3,248 (6,183) | 3,015 | 3,349 | 3,763 |
|  | 7-12 | 1,588 | 3,102 (6,048) | 2,975 | 3,571 | 4,779 |
|  | 1-12 | 4,909 | 3,775 (8,515) | 3,392 | 3,907 | 4,593 |
|  | 13-24 | 1,730 | 3,062 (5,859) | 3,067 | 3,484 | 4,285 |
| RSV-specific |  |  |  |  |  |  |
|  | 1-3 | 1,536 | 4,792 (10,809) | 4,224 | 4,692 | 5,000 |
|  | 4-6 | 1,029 | 3,397 (5,215) | 3,180 | 3,477 | 3,750 |
|  | 7-12 | 806 | 3,349 (3,850) | 3,233 | 3,838 | 5,483 |
|  | 1-12 | 3,371 | 4,021 (8,096) | 3,669 | 4,117 | 4,734 |
|  | 13-24 | 618 | 3,641 (5,804) | 3,521 | 3,921 | 4,722 |
| UBP |  |  |  |  |  |  |
|  | 1-3 | 355 | 4,503 (13,112) | 3,176 | 4,268 | 5,347 |
|  | 4-6 | 401 | 2,866 (8,153) | 2,592 | 3,019 | 3,796 |
|  | 7-12 | 782 | 2,847 (7,677) | 2,709 | 3,297 | 4,053 |
|  | 1-12 | 1,538 | 3,234 (9,348) | 2,786 | 3,449 | 4,285 |
|  | 13-24 | 1,112 | 2,741 (5,868) | 2,815 | 3,241 | 4,042 |

**Table 5.**
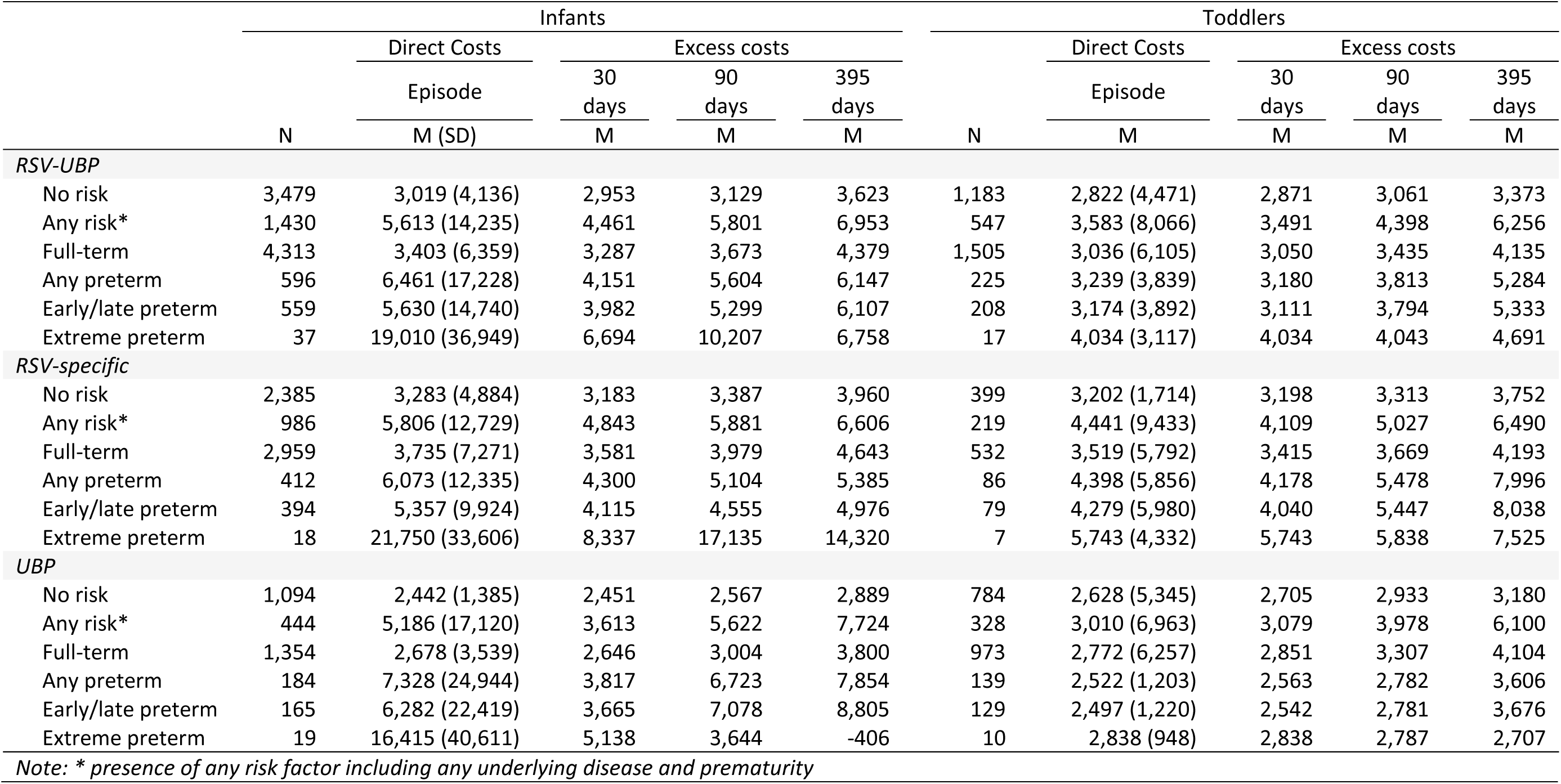
Case Numbers (N), Mean (M) and Standard Deviation (SD) of Direct Costs and Mean Excess Costs (in €) by Risk and Preterm Status in Infants and Toddlers (2014 – 2019)

Mean excess costs increased for both infants and toddlers over the follow-up periods of 30, 90 and 365 days (Table 4), whereby costs were consistently higher for patients than matched controls (Table S5). Moreover, mean excess costs were higher for children born premature or at risk (Table 5). Mean excess costs within 30 days after hospital admission were also higher for both infants and toddlers that required ICU admission during the RVS/UBP episode (11,809 € and 23,556 €), respectively) than for infants and toddlers that did not require ICU admission (3,318 € and 2,924 €, respectively). Additionally, mean excess costs within 30 days after hospital admission were higher for cases that required ventilatory support during the RVS/UBP episode, ranging from 5,808 € for oxygen therapy to 31,404 € for invasive mechanical ventilation in infants. For both infants and toddlers, cases based on an RSV episode were more costly (3,669 € and 3,521 €, respectively) than cases based on a UBP episode (2,786 € and 2,815 €, respectively).

An estimation of the annual economic burden of RSV-related hospitalizations, based on annual case numbers extrapolated to the population-level of 23,152 infants and 11,228 toddlers at mean excess costs of 3,392 € for infants and 3,067 € for toddlers, results in costs of 112,967,860 € per year for children under the age of 2 years; that is, ∼64.5 mil € for cases with an RSV-specific diagnosis (based on 15,469 cases in infants and 3,907 cases in toddlers) and ∼48.5 mil € for cases with a UBP diagnosis (based on 7,683 cases in infants and 7,321 cases in toddlers).

## Discussion

In this retrospective cohort study based on a representative sample of SHI data, we assessed a nation-wide, and population-based clinical and economic burden of RSV infections in young children < 2 years in Germany between 2014-2019, quantifying hospital incidence rate, mortality rate, HRU and associated costs of RSV. The results how that hospital incidence rate, HRU and associated costs were higher in infants than in toddlers and were highest for very young infants, infants born prematurely and infants at risk.

The results show that hospital incidence rates from 2014-2019 that were based on a wide case definition, taking RSV-UBP cases into account, (28.95-33.07 per 1,000 in infants; 10.95-18.13 per 1,000 in toddlers) are higher than previous reports for Germany from 2019-2022 that were based on a narrow case definition, considering RSV-specific cases only (14.9-28.6 per 1,000 in infants, 2.5-5.6 per 1,000 in toddlers; [11]). Though, our hospital incidence rates based on the narrow case definition are lower (18.50-23.74 per 1,000 in infants; 3.98-6.95 per 1,000 in toddlers) – likely due to the earlier observation period; the disease burden is known to increase year by year [9]. Applying an underreporting factor of 1.35 (1.3 in infants < 6 months and 1.4 in infants 6-11 months of age) as reported by Haeberer et al. [12] to our RSV-specific incidence rate results in 24.98-32.05 per 1,000 in infants which approximates estimates for the overall RSV-UBP study population. While these estimates are lower than the mean hospital incidence rate of 38.5/1,000 in infants in other industrialized countries across the world [6], they are comparable to estimates from neighboring European countries, for instance Denmark with a hospital incidence rate of 29/1,000 in infants [8]. This substantiates an estimation based on a wide case definition including RSV-UBP cases to describe the RSV-attributable clinical burden for infants and toddlers.

We also show that hospital incidence rates were highest in very young infants during the first months of life, in line with previous reports [7, 8, 25, 26]. For Germany, between 2006-2018, Johannesen et al. [26] reported substantially higher RSV-attributable hospital incidence rates of 72.5 per 1,000 in infants between 1-3 months of life than 38.6 and 17.5 per 1,000 in infants between 4-6 and 7-12 months of life, respectively. In comparison, we reported slightly lower incidence rates of 44.21, 35.36 and 20.72 per 1,000 persons in infants between 1-3, 4-6 and 7-12 months of life, respectively. However, this is likely due to the difference in methodology; while we are using counts of ICD-10 diagnosis codes, Johannesen et al. [26] modelled incidence rates using time-series estimation. The modelled estimates are comparable to average incidence rates reported across the European Union (EU): 71.5, 38.9 and 17.6 per 1,000 persons in infants between 1-3, 4-6 and 7-12 months of life, respectively [27]. As time-series modeling is less sensitive to the effects of underreporting and as the results are comparable across the EU, the burden of RSV in children may be even higher in Germany than the results of our study suggest – despite the inclusion of UBP cases during the RSV season to take underreporting into account. Future studies employing novel approaches to the estimation of incidence rates like time-series modeling are needed.

Further extending previous findings, we show in this study that the risk of contracting severe forms of RSV in newborns is approximately twice as high right after birth in infants at risk, that is infants born prematurely or with any underlying disease, compared to infants without any risk factors. Several risk factors like prematurity, bronchopulmonary dysplasia (BPD) or congenital heart disease (CHD) have been widely acknowledged as risk factors for severe outcomes of RSV, globally [28–31] and in Germany [4], but no detailed evidence is yet available on the burden of RSV in this most vulnerable population during the first few months of life in Germany. For infants with underlying disease (including BPD and CHD), we show that the risk of infection is already imminent right after birth, in the first month of life. For preterm infants, the risk is highest during the second and third month of life. As newborns have an immature immune system, and their small airways are especially affected by swelling and mucous production due to the infection and inflammation [32]), in the first few months of life infants are most vulnerable, especially when at risk (due to prematurity or any underlying disease). The results of this study show that the impact of premature birth and underlying disease on the risk for hospitalization seems to persist well beyond the first few months of life, well into toddler age and possibly beyond.

Moreover, while the hospital burden of RSV in Germany is already substantial, it represents only part of the overall burden of RSV, including largely severe cases that require hospitalization. The overall burden of RSV, however, includes also less severe RSV infections that present to the outpatient sector. The estimation of the outpatient burden is, however, challenging not least because of similar symptomology to other respiratory diseases and less established testing practice, leading to an even larger underreporting. Data on the outpatient burden are currently missing and should be assessed in the future once more advances are made to identify RSV in the outpatient sector.

HRU and associated costs underline the risk for severe outcomes of RSV in children with risk factors as seen in longer LOS and admissions to ICU. We show that for children born early/late or extreme preterm, the LOS is one and two days higher, respectively, than the previously reported 5 days [9–11]. 2.1%-8.0% infants were reported to be admitted to ICU [9–11] – largely exceeding our estimates of ∼1%, likely due to methodological differences to identify ICU admissions. The highest ICU admission rate, though, was previously found in children under 6 months of age (9.3%) [10].

Associated costs were highest in the first few months of life, especially for those at risk. This extends previous findings for Germany [9, 11] and is in line with other recent cost estimates from other European countries like France [25, 33]. For example, Kramer et al. [33] estimated mean costs per inpatient episode for full-term infants at 3,437 €, with higher costs for infants <3 months of age (3,958 €) than infants >3 months of age (3,234 €). Kramer et al. [33] also reported higher estimates in infants born prematurely at 6,324 €. For Germany, we additionally show that costs are specifically high for infants born extremely prematurely. However, even though the costs are highest for children at risk, the economic burden is driven by the large clinical burden in otherwise healthy children under the age of two years.

Both clinical and associated economic burden may be reduced in effect of a new RSV intervention program. At this point in time, three preventive measures have been granted market authorization for the prevention of RSV-LRTI in infants. One monoclonal antibody (mAB), Palivizumab [34, 35], has been used to prevent severe RSV infections in children at risk since the 1990s and is now being complemented by a recently approved mAB with a longer half-life (Nirsevimab, [36–38]). The latter has just been recommended for all infants regardless of their risk status in Germany [39]. While mAbs can protect from the time of administration, the recently approved maternal vaccine (RSVpreF, [40, 41]) provides protection immediately from birth, as children are born with protective antibodies that were transferred by the vaccinated mother during pregnancy.

### Limitations and Implications

Using health claims data to analyze clinical and economic burden has numerous strengths, but also several limitations. First, it is not possible to determine whether an RSV-specific laboratory detection assay was positive prior to ICD-10 coding, as virological confirmation is not a requirement, and the results of tests are not available in health claims data. However, RSV-codes (J12.1, J20.5, and J21.0) are usually allocated after laboratory-confirmed RSV infection in the hospital setting [9]. Second, while the database used in this study is representative of the population, available SHI databases typically include only ∼5% of the population, capturing many but likely not all cases in contrast to public databases. However, SHI databases have the advantage of more detailed information on patient characteristics like preterm or risk status which was crucial in this study to assess the burden in the most vulnerable population. Though the number of patients with such specific characteristics are low. This affects specifically results on mean costs which are most robust if the sample was large. However, to date this is the best estimation available. That is also the case for the estimation of excess costs; the best possible procedure for matching patients of such young age by at least month of life, sex and preterm status to controls was only feasible across healthcare sectors. As a result only 12% of controls included in the analysis had a hospitalization within 365 days after the index date of the matched incident patient. This was, however, the best matching procedure possible. Moreover, we employed an approach – the inclusion of UBP cases during the RSV season – that previous studies have already used in order to take underreporting due to misdiagnosis into account [e.g., 19, 20]. While overestimation may be a concern to this approach, the number of cases that are added correspond approximately to the underreporting factor that has previously been suggested for that age group [12]. Also, novel approaches like time-series modeling report even higher incidence rates compared to the standard estimation employed here [26]. However, for comparability to previous studies [9, 11], we also report estimates on a narrow case definition based on RSV-specific diagnosis codes only.

## Conclusion

This large-scale observational study is the first to provide detailed nation-wide, population-based epidemiological and economic estimates of the RSV-attributable hospital burden by months of life, preterm and risk status in Germany. The results show a substantial burden among the youngest children with RSV infections, right after birth. Although the highest risk of severe RSV was found in children born prematurely or with an underlying disease, most children who needed to be hospitalized were not at risk – emphasizing the need for broad protection already at birth.

## Data Availability

All data produced in the present study are available upon reasonable request to the authors.

## Acknowledgements

Medical writing support was provided by Qi Yan, PhD, MS (Pfizer, Inc). We thank Julia Schiffner-Rohe for valuable discussions throughout the study process.

