## Supplements for "Clinical and economic inpatient burden of respiratory syncytial virus (RSV) infections in children < 2 years of age in Germany, 2014-2019: a retrospective health claims analysis"

Authors in order:

Caroline Lade[1] – Lead and corresponding author

Lea Bayer[1]

Bennet Huebbe[2]

Jennifer Riedel[3]

Sima Melnik[3]

Gordon Brestrich[4]

Christof von Eiff[1]

Tobias Tenenbaum[5]

Affiliation:

1 Pfizer Pharma GmbH, Berlin, Germany

2 IGES Institute, Berlin, Germany

3 Gesundheitsforen Leipzig GmbH, Leipzig, Germany

4 Pfizer Inc, New York, USA

5 Sana Klinikum Lichtenberg, Clinic for Child and Adolescent Medicine, Academic Teaching Hospital Charité-Universitätsmedizin Berlin, Germany

**Supplementary Materials**

**Table S1** RSV seasons according to Cai et al., 2022

| Season | Calendar weeks |
|---|---|
| 2013/2014 | 3-19 |
| 2014/2015 | 46-12 |
| 2015/2016 | 49-15 |
| 2016/2017 | 44-11 |
| 2017/2018 | 50-14 |
| 2018/2019 | 49-12 |
| 2019/2020 | 50-14 |

**Table S2** Risk Factor Definition by ICD-10 Codes

| Risk Condition | ICD-10-GM Code | Description |
|---|---|---|
| Conditions originating in the perinatal period | P07 | Disorders of newborn related to short gestation and low birth weight, not elsewhere classified |
| | P23 | Congenital pneumonia |
| | P25 | Interstitial emphysema and related conditions originating in the perinatal period |
| | P27.1 | Bronchopulmonary dysplasia originating in the perinatal period |
| | P27.8 | Other chronic respiratory diseases originating in the perinatal period |
| | P28 | Other respiratory conditions originating in the perinatal period |
| | P27.9 | Unspecified chronic respiratory disease originating in the perinatal period |
| | P29 | Cardiovascular disorders originating in the perinatal period |
| Chronic cardiac disease | Q20-26 | Congenital malformations of the circulatory system |
| | I42, I43, I50-I52 | Other forms of heart disease |
| Chronic respiratory disease | J43 | Chronic lower respiratory diseases |
| | J96.1 | Chronic respiratory failure |
| | E84 | Cystic fibrosis |
| | Q79.0, Q79.1 | Congenital diaphragmatic hernia, Other congenital malformations of diaphragm |
| | Q30 - Q34 | Congenital malformations |
| Immunosuppression | D57 | Sickle-cell disorders |
| | D80, D81, D83, D84, | Certain disorders involving the immune mechanism |
| | Q90-Q95 | Chromosomal abnormalities, not elsewhere classified |
| | Z94.0-Z94.4 | Transplanted organ and tissue status |
| Neurological disease | G12, G13 | Systemic atrophies primarily affecting the central nervous system |
| | G31.8, G31.9, G32 | Other degenerative diseases of the nervous system |
| | G40 | Epilepsy |
| | G71 | Primary disorders of muscles |
| | G93.1, G93.4 | Other disorders of the nervous system |
| | P91 | Other disturbances of cerebral status of newborn |
| Patient History | Z29.1 | Encounter for prophylactic immunotherapy |
| Diabetes mellitus | E10 | Type 1 diabetes mellitus |
| | P70.2 | Neonatal diabetes mellitus |

**Table S3** Proportion (%) of inpatient cases per calendar months in infants and toddlers across 2014-2019

| | **Infants** | | | **Toddlers** | | |
|---|---|---|---|---|---|---|
| Calendar months | RSV-UBP N = 5,565 | RSV-specific N = 3,706 | UBP N = 1,859 | RSV-UBP N = 2,131 | RSV-specific N = 741 | UBP N = 1,390 |
| **January** | 24.0 | 24.6 | 23.0 | 26.9 | 26.3 | 27.2 |
| **February** | 28.0 | 28.5 | 26.9 | 29.8 | 32.0 | 28.7 |
| **March** | 19.0 | 19.1 | 18.7 | 17.7 | 19.0 | 17.1 |
| **April** | 5.9 | 6.5 | 4.6 | 2.1 | 4.3 | 0.9 |
| **May** | 1.3 | 1.9 | - | 0.5 | 1.5 | - |
| **June** | 0.3 | 0.4 | - | - | - | - |
| **July** | - | - | - | - | - | - |
| **August** | 0.2 | 0.2 | - | - | - | - |
| **September** | 0.1 | 0.2 | - | - | - | - |
| **October** | 0.5 | 0.6 | - | 0.5 | 1.1 | - |
| **November** | 3.5 | 2.9 | 4.7 | 3.6 | 2.2 | 4.3 |
| **December** | 17.3 | 15.0 | 21.8 | 18.6 | 12.8 | 21.7 |

Note: UBP cases were only taken into consideration within the RSV season (see Table S1)

**Table S4** Case Numbers (N) and Incidence Rates (IR) per 1,000 person-years per Month of Life by Pre-term Status and Underlying Disease across 2014-2019

| **Months of life** | **All** | | **Full Term** | | **Pre-term** | | **Underlying disease** | |
|---|---|---|---|---|---|---|---|---|
| | N | IR | N | IR | N | IR | N | IR |
| ***RSV-UBP*** | | | | | | | | |
| **1** | 467 | 30.46 | 420 | 29.21 | 47 | 49.36 | 163 | 110.16 |
| **2** | 878 | 57.28 | 785 | 54.60 | 93 | 97.66 | 126 | 85.15 |
| **3** | 688 | 44.88 | 590 | 41.04 | 98 | 102.91 | 101 | 68.26 |
| **4** | 579 | 37.77 | 516 | 35.89 | 63 | 66.16 | 85 | 57.45 |
| **5** | 559 | 36.47 | 482 | 33.53 | 77 | 80.86 | 84 | 56.77 |
| **6** | 488 | 31.83 | 419 | 29.14 | 69 | 72.46 | 68 | 45.96 |
| **7** | 441 | 28.77 | 378 | 26.29 | 63 | 66.16 | 78 | 52.71 |
| **8** | 381 | 24.85 | 319 | 22.19 | 62 | 65.11 | 70 | 47.31 |
| **9** | 288 | 18.79 | 234 | 16.28 | 54 | 56.71 | 61 | 41.23 |
| **10** | 264 | 17.22 | 232 | 16.14 | 32 | 33.60 | 45 | 30.41 |
| **11** | 266 | 17.35 | 229 | 15.93 | 37 | 38.86 | 47 | 31.76 |
| **12** | 266 | 17.35 | 221 | 15.37 | 45 | 47.26 | 46 | 31.09 |
| **13** | 206 | 16.84 | 179 | 15.61 | 27 | 35.36 | 46 | 33.06 |
| **14** | 206 | 16.84 | 174 | 15.18 | 32 | 41.91 | 43 | 30.90 |
| **15** | 195 | 15.95 | 164 | 14.30 | 31 | 40.60 | 34 | 24.43 |
| **16** | 206 | 16.84 | 180 | 15.70 | 26 | 34.05 | 39 | 28.03 |
| **17** | 233 | 19.05 | 200 | 17.44 | 33 | 43.22 | 52 | 37.37 |
| **18** | 200 | 16.35 | 173 | 15.09 | 27 | 35.36 | 47 | 33.78 |
| **19** | 187 | 15.29 | 157 | 13.69 | 30 | 39.29 | 41 | 29.46 |
| **20** | 174 | 14.23 | 148 | 12.91 | 26 | 34.05 | 32 | 23.00 |
| **21** | 151 | 12.35 | 127 | 11.08 | 24 | 31.43 | 28 | 20.12 |
| **22** | 147 | 12.02 | 121 | 10.55 | 26 | 34.05 | 29 | 20.84 |
| **23** | 108 | 8.83 | 91 | 7.94 | 17 | 22.27 | 18 | 12.94 |
| **24** | 118 | 9.65 | 101 | 8.81 | 17 | 22.27 | 23 | 16.53 |
| ***RSV**-specific* | | | | | | | | |
| **1** | 408 | 26.62 | 369 | 25.67 | 39 | 40.96 | 144 | 97.32 |
| **2** | 693 | 45.21 | 624 | 43.40 | 69 | 72.46 | 97 | 65.56 |
| **3** | 534 | 34.84 | 456 | 31.72 | 78 | 81.91 | 70 | 47.31 |
| **4** | 421 | 27.46 | 374 | 26.01 | 47 | 49.36 | 62 | 41.90 |
| **5** | 381 | 24.85 | 331 | 23.02 | 50 | 52.51 | 56 | 37.85 |
| **6** | 330 | 21.53 | 282 | 19.61 | 48 | 50.41 | 47 | 31.76 |
| **7** | 246 | 16.05 | 208 | 14.47 | 38 | 39.91 | 37 | 25.01 |
| **8** | 194 | 12.66 | 155 | 10.78 | 39 | 40.96 | 36 | 24.33 |
| **9** | 144 | 9.39 | 116 | 8.07 | 28 | 29.40 | 32 | 21.63 |
| **10** | 121 | 7.89 | 103 | 7.16 | 18 | 18.90 | 17 | 11.49 |
| **11** | 126 | 8.22 | 112 | 7.79 | 14 | 14.70 | 27 | 18.25 |
| **12** | 108 | 7.05 | 93 | 6.47 | 15 | 15.75 | 21 | 14.19 |
| **13** | 96 | 7.85 | 83 | 7.24 | 13 | 17.03 | 24 | 17.25 |
| **14** | 79 | 6.46 | 65 | 5.67 | 14 | 18.34 | 15 | 10.78 |
| **15** | 57 | 4.66 | 47 | 4.10 | 10 | 13.10 | 13 | 9.34 |
| **16** | 71 | 5.81 | 60 | 5.23 | 11 | 14.41 | 13 | 9.34 |
| **17** | 76 | 6.21 | 65 | 5.67 | 11 | 14.41 | 19 | 13.65 |
| **18** | 64 | 5.23 | 56 | 4.88 | 8 | 10.48 | 14 | 10.06 |
| **19** | 65 | 5.32 | 53 | 4.62 | 12 | 15.72 | 18 | 12.94 |
| **20** | 65 | 5.32 | 54 | 4.71 | 11 | 14.41 | 11 | 7.91 |
| **21** | 50 | 4.09 | 42 | 3.66 | 8 | 10.48 | 11 | 7.91 |
| **22** | 54 | 4.42 | 44 | 3.84 | 10 | 13.10 | 12 | 8.62 |
| **23** | 33 | 2.70 | 27 | 2.35 | 6 | 7.86 | 6 | 4.31 |
| **24** | 31 | 2.53 | 29 | 2.53 | <5 | NA | 10 | 7.19 |

**Table S5** Case Numbers (N), Mean (M) and Standard Deviation (SD) of Costs in RSV-UBP-, RSV-specific and UBP-patients in Infants and Toddlers

| | | | Controls | | | Incident Cases | | |
|---|---|---|---|---|---|---|---|---|
| | Month of life | N | 30 days M (SD) | 90 days M (SD) | 365 days M (SD) | 30 days M(SD) | 90 days M(SD) | 365 days M(SD) |
| *RSV-UBP* | | | | | | | | |
| | 1-3 | 1,891 | 201 (1,695) | 413 (3,531) | 819 (6,766) | 4,228 (6,552) | 5,025 (9,789) | 5,884 (13,951) |
| | 4-6 | 1,430 | 99 (1,538) | 188 (2,098) | 480 (2,876) | 3,114 (3,238) | 3,537 (6,796) | 4,243 (8,231) |
| | 7-12 | 1,588 | 32 (355) | 92 (641) | 324 (2,909) | 3,007 (2,689) | 3,663 (8,429) | 5,103 (24,383) |
| | 1-12 | 4,909 | 116 (1,357) | 243 (2,497) | 560 (4,777) | 3,508 (4,717) | 4,151 (8,591) | 5,153 (16,952) |
| | 13-24 | 1,730 | 14 (176) | 65 (461) | 223 (989) | 3,081 (5,606) | 3,549 (9,166) | 4,507 (15,131) |
| *RSV-specific* | | | | | | | | |
| | 1-3 | 1,536 | 196 (1,624) | 335 (2,487) | 699 (4,743) | 4,420 (6,963) | 5,027 (9,427) | 5,699 (12,334) |
| | 4-6 | 1,029 | 117 (1,786) | 205 (2,226) | 505 (3,010) | 3,298 (3,352) | 3,682 (6,091) | 4,255 (7,005) |
| | 7-12 | 806 | 38 (416) | 99 (681) | 252 (1,161) | 3,271 (2,061) | 3,937 (6,570) | 5,735 (32,098) |
| | 1-12 | 3,371 | 134 (1,490) | 239 (2,109) | 533 (3,656) | 3,803 (5,181) | 4,356 (7,905) | 5,267 (18,188) |
| | 13-24 | 618 | 20 (207) | 64 (394) | 166 (808) | 3,541 (3,704) | 3,985 (6,638) | 4,889 (9,354) |
| *UBP* | | | | | | | | |
| | 1-3 | 355 | 221 (1,976) | 750 (6,293) | 1,336 (12,105) | 3,397 (4,255) | 5,018 (11,235) | 6,684 (19,461) |
| | 4-6 | 401 | 50 (498) | 145 (1,726) | 416 (2,501) | 2,642 (2,877) | 3,164 (8,336) | 4,212 (10,766) |
| | 7-12 | 782 | 25 (280) | 84 (597) | 398 (3,975) | 2,735 (3,189) | 3,381 (9,985) | 4,452 (12,057) |
| | 1-12 | 1,538 | 77 (1,005) | 254 (3,186) | 620 (6,600) | 2,863 (3,403) | 3,702 (9,917) | 4,905 (13,865) |
| | 13-24 | 1,112 | 10 (156) | 65 (494) | 254 (1,075) | 2,825 (6,411) | 3,307 (10,302 | 4,295 (17,538) |

**Table S6** Overall base population and extrapolation factors for infants and toddlers per calendar year between 2014-2019

| | Live birth in Germany | DADB infants | Extrapolation Factor infants | DADB toddler | Extrapolation Factor toddlers |
|---|---|---|---|---|---|
| 2014 | 714,927 | 30,605 | 23.36 | NA | NA |
| 2015 | 737,575 | 30,211 | 24.41 | 28,904 | 25.52 |
| 2016 | 792,141 | 30,853 | 25.67 | 28,731 | 27.57 |
| 2017 | 784,901 | 31,609 | 24.83 | 29,508 | 26.60 |
| 2018 | 787,523 | 31,191 | 25.25 | 30,272 | 26.01 |
| 2019 | 778,090 | 29,483 | 26.39 | 29,337 | 26.52 |

**Figure S1** Patient flow

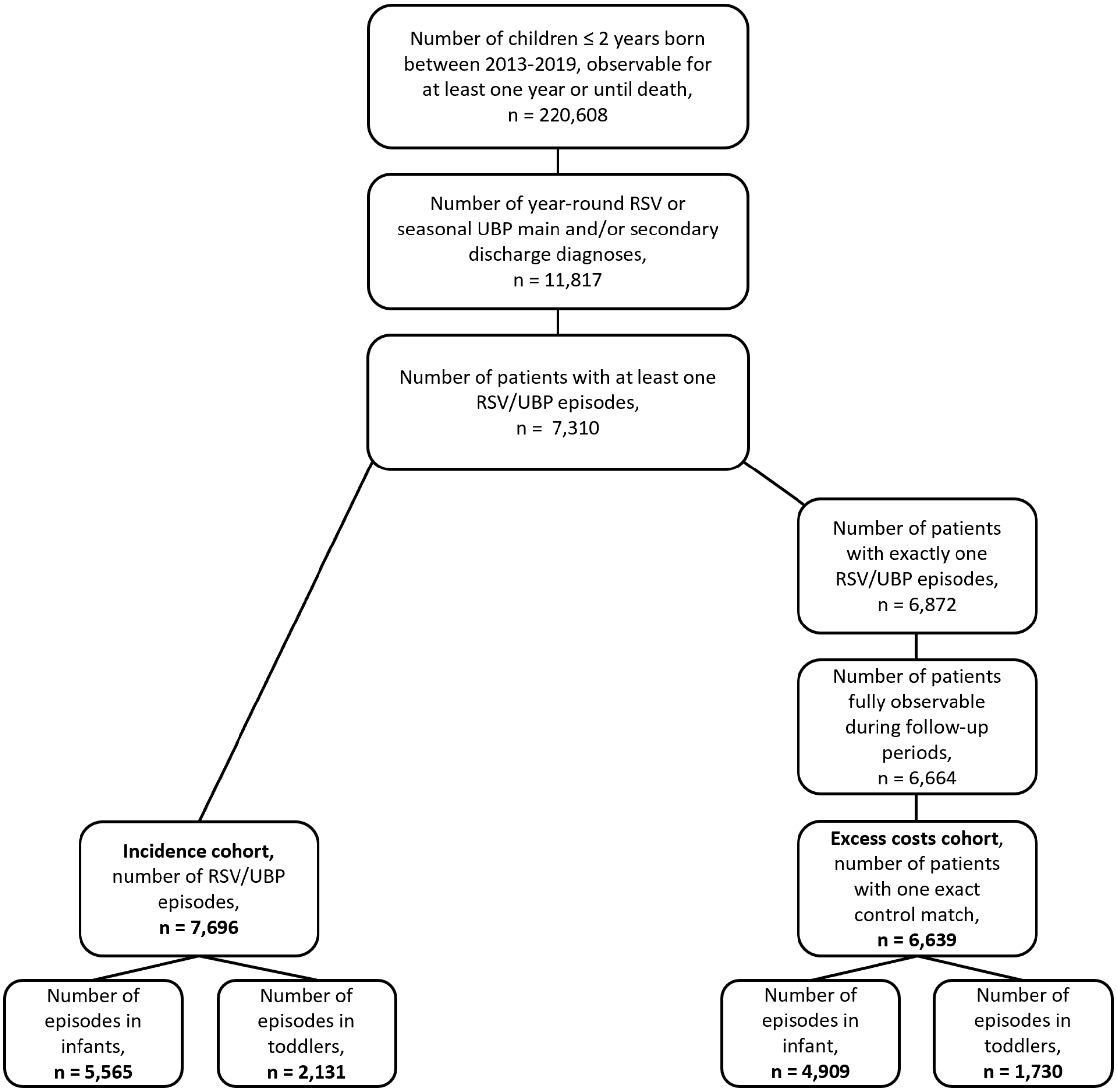
